# Development and validation of an algorithm to identify front-line clinicians using EHR audit log data

**DOI:** 10.64898/2026.02.13.26346268

**Authors:** Laura R. Baratta, Joanne Wang, Bailey W. Osweiler, Daphne Lew, Elise Eiden, Thomas Kannampallil, Sunny S. Lou

**Author notes:** Corresponding author:* Sunny S Lou, Department of Anesthesiology, 660 S Euclid Ave, Box 8054, St Louis, MO 63110.

## Abstract

**Background:** Interprofessional teams are central to high quality patient care. However, identifying the clinician primarily responsible for a patient requires labor-intensive methodologies. Although electronic health record (EHR) audit logs offer a scalable alternative, its use for identifying frontline clinicians is underdeveloped.

**Objective:** To develop and validate an algorithm utilizing EHR audit logs to identify the primary frontline clinician per patient day of an encounter and to describe care continuity patterns.

**Method:** This was a cross-sectional cohort study of adult inpatient medicine encounters at 12 hospitals in a single health system using a shared EHR. Admissions from February 1, 2023–April 30, 2023, with length of stay of at least 3 days and without an intensive care unit admission were included. Four algorithm iterations were designed to identify the attending physician, resident, or advanced practice provider primarily responsible for patient care on each patient-day. Performance of each algorithm was compared with manual chart review on 1,401 patient-days from 246 randomly sampled patient encounters. Accuracy between an algorithm and the chart review standard was compared using McNemar’s test with Bonferroni adjusted p-values.

**Results:** The best performing algorithm correctly identified the primary clinician responsible for patient care on 91% of patient-days (1,268/1,401), outperforming the naïve approach using frequency of actions (78% accuracy, 1,098/1,401, p<0.001). Algorithm errors were attributable to misidentified specialty and ambiguity on days with transitions of care or shared responsibilities between clinicians. The best performing algorithm was applied to the entire cohort (5,801 encounters and 34,001 patient-days) where it identified attending physicians, resident physicians, and APPs as the frontline clinician for 26,750 (79%), 3,106 (9%), and 4,145 (12%) of patient days respectively. Each encounter had a median of 1 (IQR 0-2) handoff between frontline clinicians.

**Conclusions:** We developed a scalable, audit log-based algorithm to determine the front-line clinician with excellent accuracy compared with manual chart review.

## Background and Significance

Inpatient care is delivered by interprofessional teams whose composition and responsibilities frequently change over the course of a hospitalization (1, 2). Within these teams, identifying the clinician primarily responsible for a patient’s day-to-day care (i.e., the primary frontline clinician) is often ambiguous (3). This ambiguity is particularly pronounced in academic and team-based medicine services, where attending physicians, resident physicians, and advanced practice practitioners (APPs) may share or transition responsibility across patient-days. Accurately identifying the primary frontline clinician is essential for measuring clinician-level quality, assessing continuity of care, evaluating handoffs, and attributing accountability for patient outcomes (4–7).

Historically, characterizing care teams and identifying responsible clinicians has relied on labor-intensive or resource-intensive methods such as direct observation, surveys, or manual chart review (8–14). While these approaches can be accurate, they are costly, difficult to scale, and impractical for large health system-level analyses. Claims-based attribution methods offer scalability but are typically limited to identifying the single billing provider for an encounter or hospitalization, usually the attending physician, and do not reliably capture resident or APP participation or day-to-day rotating shifts in responsibility (3, 15–19).

The widespread adoption of electronic health records (EHRs) has created new opportunities to address these limitations. EHR audit logs capture detailed, time-stamped records of clinician interactions with the patient chart, including actions that display, review, or modify clinical data. Because much of inpatient clinical work is mediated through the EHR, audit logs provide a rich and scalable data source that reflects real-world care delivery processes (20, 21). Prior informatics research has demonstrated the feasibility of using audit logs to infer care team membership, clinician workload, and patterns of clinical activity (20, 22).

However, existing audit log–based approaches have important limitations for clinician attribution. Many rely on a narrow or context-specific set of EHR actions tailored to particular roles, workflows, or clinical environments, limiting generalizability across hospitals and services (2, 15). Few studies have rigorously validated audit log–based attribution against a gold standard such as manual chart review, particularly at the patient-day level and across multiple clinician roles.

Beyond attribution, accurately identifying the primary frontline clinician enables the study of care continuity and handoffs, which are known to influence patient safety and outcomes (22–24). Discontinuities in care and frequent handoffs have been associated with communication failures, medical errors, and adverse events, while greater continuity has been linked to improved outcomes and patient satisfaction (25–28). Despite this, continuity is difficult to measure at scale in inpatient settings due to the lack of reliable, granular attribution of responsibility over time.

### Objectives

In this study, we developed and validated a generalizable algorithm that uses EHR audit log data to identify the primary frontline clinician—attending physician, resident physician, or APP—for each patient-day during inpatient medicine encounters. We evaluated algorithm performance against manual chart review and applied the algorithm at scale to characterize patterns of clinician continuity and handoffs across hospitalizations.

## Methods

### Study Setting

This was a cross-sectional cohort study conducted at 12 inpatient hospitals affiliated with BJC Health and Washington University School of Medicine. Hospitals included both academic and community practice settings serving diverse rural, suburban, and urban populations across Missouri and Illinois. All hospitals used the same Epic EHR system (Epic Systems, Verona, WI).

### Study Sample

All adult inpatient medicine encounters occurring between February 1, 2023, through April 30, 2023, were included. This consisted of hospitalizations for patients arriving through the emergency department (ED) or directly to an inpatient medicine clinical service. Additional encounter inclusion criteria included if (1) they were admitted to an inpatient medicine service throughout their stay, (2) had a length of stay of at least 72 hours, and (3) patient’s age >= 18.

Encounters were excluded if (1) the patient was transferred to the ICU at any point during their stay, (2) if they were admitted to a children’s hospital or (3) if no audit log actions were observed between admission and discharge or after ED transfer.

To construct the study cohort, admission/discharge/transfer (ADT) data was utilized to identify adult inpatient medicine encounters whose admission and discharge fell within the study period. The ADT table was used to identify the admission time, discharge time, transfer time out of ED, and whether there was ICU level of care.

A CONSORT-style diagram illustrating cohort construction is shown in Figure 1.

**Figure 1:**
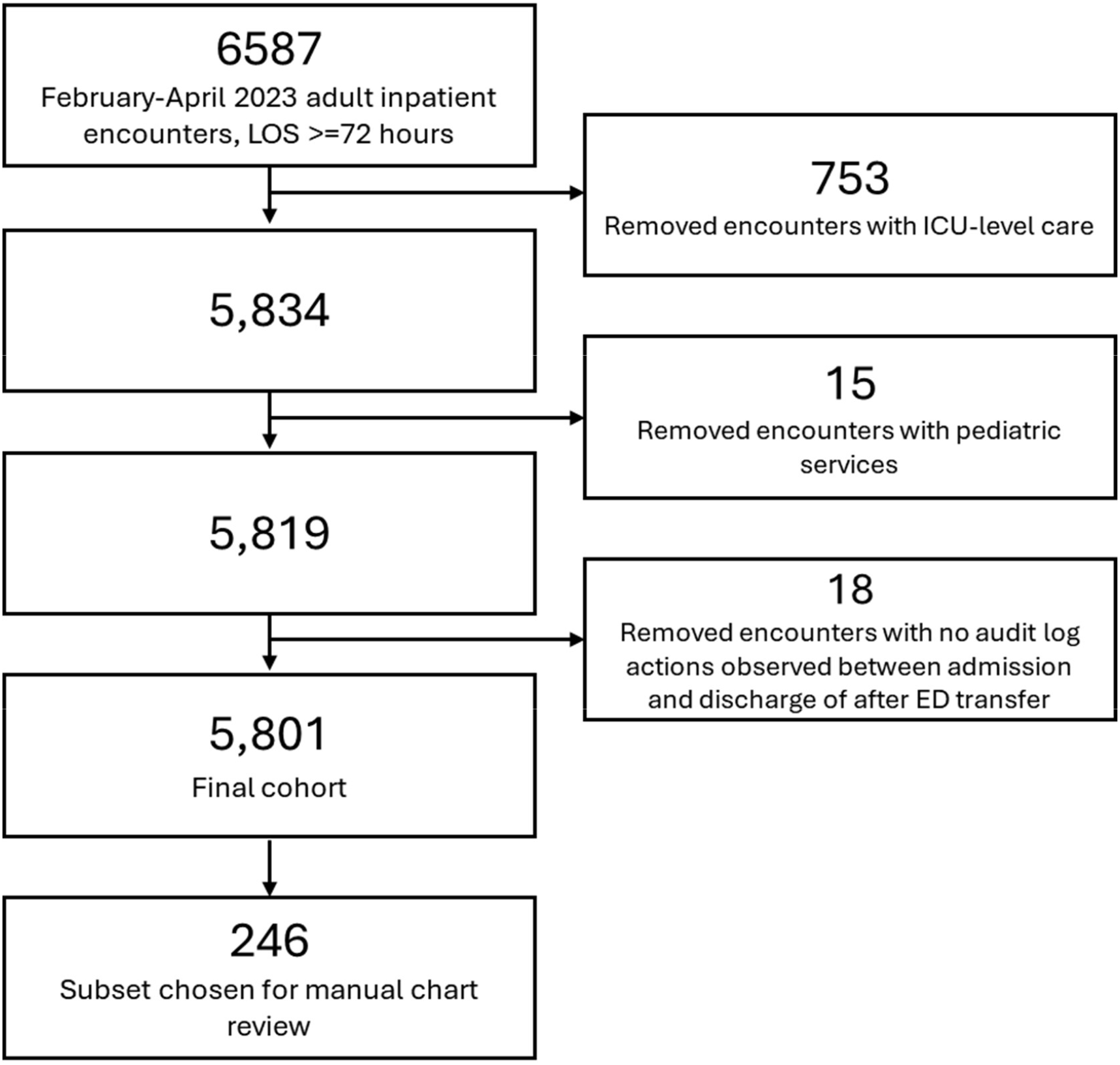
CONSORT-style diagram describing cohort identification and selection. The diagram summarizes the initial inclusion and subsequent exclusion criteria along with the number of encounters removed at each step, yielding the final cohort. LOS – length of stay, ED – emergency department, ICU – intensive care unit.

### Data sources and preprocessing

The following data sources were extracted from Epic EHR’s Clarity database: EHR audit logs from the ACCESS_LOG table, EHR login information used to determine the user’s specialty or department, and administrative and demographic information on individual clinicians. The unit of analysis was the patient-day (i.e., a calendar day), and all audit log actions performed on that patient’s chart on each patient-day of an encounter were considered in the analysis. Any actions performed in the ED were excluded from analysis, as our goal was to determine the primary inpatient medicine frontline clinician. Only actions performed by attending physicians, resident physicians, or APPs were included.

### Algorithm design

Our primary goal was to develop an algorithm to identify the primary frontline clinician responsible for a patient’s care on each day of their inpatient stay (i.e., per patient-day). All algorithms used EHR audit log activities to select the primary clinician per patient day.

Four variations of the algorithm were developed (Figure 2):

**Figure 2:**
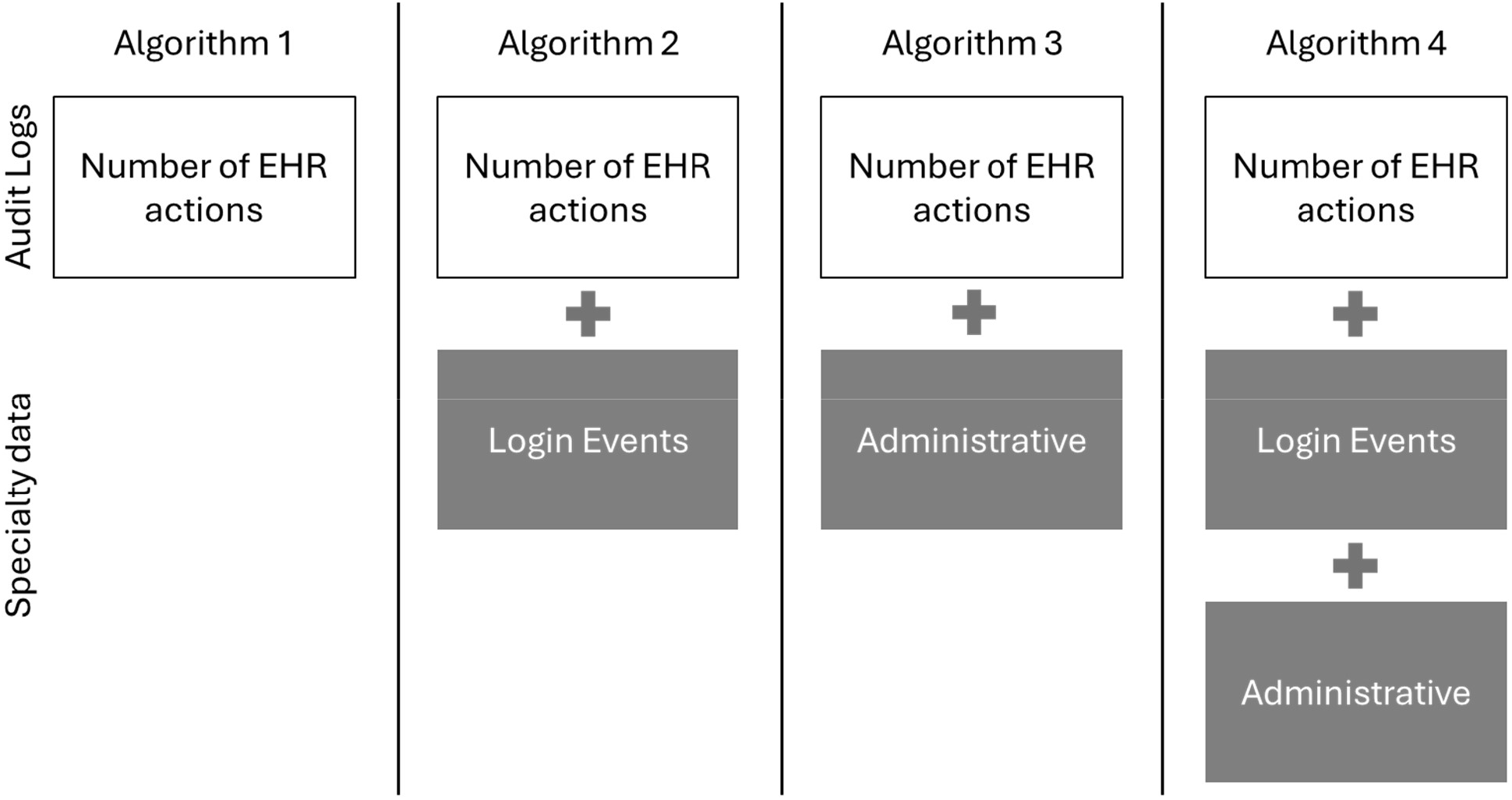
Graphical representation of each algorithm variant and the data sources utilized to identify the primary clinician.

#### 1) Select the clinician with the most EHR actions

Our initial assumption was that the primary clinician responsible for patient care would perform the greatest number of audit log actions on that patient’s chart, as they are expected to be the clinician who places the orders and reviews the patient’s chart in greatest detail. Audit logs have previously been shown to be a proxy for clinical workload (29, 30), and we anticipated that the primary clinician would perform the greatest amount of EHR work for the patient. This is consistent with prior work demonstrating that the primary clinicians of a given patient spend substantially more time in that patient’s chart (15). Therefore, the initial version of our algorithm simply selected the clinician who performed the greatest number of audit log actions on that patient for that day as the primary clinician.

#### 2) Medicine defined by login department

Several refinements to our initial algorithm were introduced to assess whether accuracy could be improved by introducing additional constraints on the specialty of the clinicians eligible to be selected as the frontline clinician.

Algorithm 2 was restricted to only select clinicians whose login department matched a medicine assignment. Each time a user logs into the Epic EHR, they must select a “Login Department”, which determines the user interface and navigation panes presented to the user. These login departments are customized for each clinical service assignment; therefore, we restricted the algorithm to only choose from clinicians whose most frequent login department on that day was “Hospitalist” or “Internal Medicine”. If no clinician was found from this criterion, we reverted to the physician performing the most actions for that patient day regardless of specialty assignment.

#### 3) Medicine defined by administrative data

Algorithm 3 used an alternative source of specialty data from administrative records. These specialty data were associated with individual clinician records in the EHR, are self-reported by individual departments, and are used for business reporting purposes. Algorithm 3 preferentially selected among the clinicians whose administrative specialty matched “Hospitalist” or “Internal Medicine”. If no clinician was found from this criterion, we reverted to the physician performing the most actions for that patient day regardless of specialty assignment.

#### 4) Hybrid approach

Algorithm 4 used both specialty data sources to improve specialty classification. We defined a tiered eligibility mask that selected clinicians in the following order (1) if login department indicated a medicine service assignment as previously described above or (2) if administrative specialty records indicated a medicine specialty as previously described above. Therefore, this hybrid approach preferentially retained the top-ranked clinician by total actions per patient-day among those meeting this eligibility mask. If no clinician was selected by these criteria, we fell back to select the physician performing the most actions for that patient-day regardless of specialty assignment.

The goal of utilizing both data sources was to compensate for nonstandard user preferences for “Login Department” or administrative errors in capturing specialty information. Therefore, this hybrid method was optimized to use concordant information from both datasets when available, and to fall back on the most reliable source when discrepancies arose, with the goal of maximizing the accuracy of specialty identification for the primary clinician.

### Chart Review

We validated the algorithms using manual chart review. 246 encounters were randomly sampled from the study period (Table 1). Three independent reviewers with clinical expertise (1 attending physician SSL, 2 medical students JW LRB) conducted the chart review, with overlapping and individual subsets of encounters assigned to each reviewer.

**Table 1.** Characteristics of the study cohort, including admission location, time spent in the emergency department (ED) (hours), inpatient length of stay (hours), patient demographics, and top 10 diagnosis-related group (DRG) names and codes. Continuous variables are summarized as median (IQR) and categorical variables as percentage (n/N); unless otherwise indicated.

| **Characteristic** | **Distribution (N = 5801 encounters)** |
| --- | --- |
| **Admission Location** |  |
| Emergency Department (ED) | 96% (5543/5801) |
| Direct to Inpatient Floor | 4% (258/5801) |
| **Hospital type** |  |
| Academic | 1118 (19%) |
| Community | 4683 (81%) |
| Time in ED (hours) | 8 (5-12) |
| Length of Stay (hours) | 171 (117-269) |
| Patient Age | 68 (56-79) |
| **Sex** |  |
| Female | 55% (3195/5801) |
| Male | 45% (2606/5801) |
| **Insurance** |  |
| Medicare | 67% (3890/5801) |
| Medicaid | 16% (905/5801) |
| Dual Medicare Medicaid | 0.05% (3/5801) |
| Private | 14% (831/5801) |
| Other | 2% (106/5801) |
| Unknown | 1% (66/5801) |
| **Marital Status** |  |
| Married | 35% (2030/5801) |
| Divorce/separated | 15% (851/5801) |
| Single | 28% (1688/5801) |
| Widowed | 20% (1150/5801) |
| Unknown | 1% (82/5801) |
| **Race** |  |
| White | 65% (3782/5801) |
| Black or African American | 33% (1914/5801) |
| Other | 1% (81/5801) |
| Unknown | 0.4% (24/5801) |
| **10 Most Commonly Observed Diagnosis Related Groups (DRG code)** |  |
| Septicemia or severe sepsis without MV > 96 hours with MCC (871) | 6% (360/5801) |
| Heart failure and shock with MCC (291) | 6% (320/5801) |
| Respiratory infections and inflammations with MCC (177) | 3% (194/5801) |
| Esophagitis, gastroenteritis and miscellaneous digestive disorders without MCC (392) | 2% (139/5801) |
| Simple pneumonia and pleurisy with MCC (193) | 2% (134/5801) |
| Cellulitis without MCC (603) | 2% (127/5801) |
| Septicemia or severe sepsis without MV >96 hours without MCC (872) | 2% (126/5801) |
| Kidney and urinary tract infections without MCC (696) | 2% (107/5801) |
| Renal failure with CC (683) | 2% (106/5801) |
| Renal failure with MCC (686) | 2% (100/5801) |

For each selected encounter, the reviewers accessed the patient’s chart and reviewed clinical notes for each patient-day. Reviewers were instructed to read the clinical notes to identify which clinician was most likely to have been the primary frontline clinician on each day. Reviewers were also provided with a list of the 5 attending physicians, resident physicians and APPs who had made the most audit log actions on the patient’s chart that day; this was provided for reference, but the reviewers were not required to use this information.

Among the overlapping set of 35 encounters that all 3 reviewers assessed, the interrater reliability was 0.94 by Fleiss’ kappa, indicating excellent agreement.

### Statistical Analysis

We summarized patient-, encounter-, and clinician-level characteristics using simple descriptive statistics. Categorical variables were reported as counts and percentages. Continuous variables were reported using median and interquartile range.

For each algorithm configuration, we calculated the proportion of patient-days in which the algorithm-identified primary clinician exactly matched the chart review standard. When comparing accuracy between algorithm configurations, we used McNemar’s test for paired binary outcomes and p-values were adjusted via the Bonferroni method to account for multiple comparisons.

The best performing algorithm was then applied to the full cohort to assign a primary clinician for each patient-day. Using these assignments, we also determined the supervision structure across encounters by determining the fraction of encounters that involved attending physicians alone vs team-based care patterns involving attending physicians, APPs, and resident physicians. Finally, we quantified handoffs as the number of times the primary clinician changed between consecutive patient-days.

All statistical tests were two-sided, with p < 0.05 considered statistically significant. The primary clinician identification algorithm was implemented using Python 3.9.7. Statistical analyses were conducted using R version 4.2.2. Code can be found at: https://github.com/thomas-k-wustl/Front-line-clinician-algorithm

## Results

### Cohort statistics

This study included 5801 encounters over the study period. 96% of encounters originated through the ED (5543/5801). Patients spent a median of 8 hours (IQR: 5-12) in the ED before being transferred to an inpatient medicine floor (Table 1). The median length of stay was 7 (IQR: 4-11) days.

The study cohort was 55% (3195/5801) women, and 65% white (3792/5801), 33% black (1914/5801), 1% other (81/5801), and 0.4% unknown (24/5801). Most patients had Medicare insurance (3890/5801, 67%). The most common Diagnosis Related Groups (DRG code) were Septicemia or severe sepsis without MV > 96 hours with MCC (871), Heart failure and shock with MCC (291), and Respiratory infections and inflammations with MCC (177) (Table 2).

**Table 2.** Accuracy of algorithms to manual chart review along with p-values for comparison of each algorithm to Algorithm 1 as reference. Bonferroni adjustment was used for multiple comparisons.

| Algorithm | Accuracy | P value vs Algorithm 1 |
| --- | --- | --- |
| 1 | 78% (1098/1401) | Reference |
| 2 | 89% (1242/1401) | 3.4e-17 |
| 3 | 83% (1158/1401) | 9.1e-3 |
| 4 | 91% (1268/1401) | 6.3e-27 |

### Validation of algorithm performance

246 encounters were randomly sampled for chart review. Algorithm 1, which simply selected the clinician with the most audit log actions on each day, correctly identified the primary frontline clinician on 78% of patient-days (1098/1401) (Table 2). Algorithm 2, which restricted the algorithm to choose internal medicine physicians based on login specialty, had an accuracy of 89% (1242/1401, p < 0.001 compared to Algorithm 1). Algorithm 3, which restricted the algorithm’s choice using administrative specialty data had an accuracy of 83% (1158/1401 p=9.1e-3 compared to algorithm 1). Algorithm 4, the hybrid approach, had the highest performance with an accuracy of 91% (1268/1401 p < 0.001 compared to algorithm 1, see Supplemental Table 2 for pairwise comparisons).

Errors for Algorithm 4 were manually reviewed and fell into 3 categories: (1) incorrect specialty information (88/133, 66%) (e.g., the true frontline clinician was miscategorized as a non–internal medicine specialty or a consulting clinician was miscategorized as internal medicine); (2) handover-related errors (24/133, 18%) (e.g., where the nighttime admitting physician and daytime physicians both performed actions on a patient’s chart on the same calendar day) – in these cases, it was not clear which should be considered the primary front-line clinician; (3) collaborative team care (21/133, 16%), where the division of tasks among team members appeared to be jointly managed across multiple clinicians creating ambiguity in the primary clinician (e.g., a resident and intern physician or a hospitalist and APP sharing a patient). These results suggest that most errors were related to poor quality source specialty information, although the algorithm also struggled in collaborative care scenarios where the primary clinician may be difficult for humans to identify as well.

### Structure of inpatient medicine encounters

We then applied Algorithm 4 to the entire study cohort to evaluate supervision structures and continuity of care. Attending physicians, resident physicians, and APPs were the frontline clinician for 26,750 (79%), 3,106 (9%), and 4,145 (12%) of patient-days, respectively. 68% (3965/5801) of encounters involved only attending physicians as frontline clinicians throughout the entire stay, 20% (1172/5801) involved attending physicians and APPs, and 11% (640/5801) involved attending physicians and resident physicians (Table 3, Supplemental Table 3).

**Table 3.** Distribution of medicine care team structures as identified by Algorithm 4 across the study cohort. Data is presented as total counts (%), median (IQR) actions per day, and fraction of days (%) each role performed audit log actions.

| **Supervision Structure** | **Number of Encounters** | **EHR Actions per Day** | **Fraction of Days with Each Role as Front-Line** |
| --- | --- | --- | --- |
| Attending physician alone | 3965 (68%) | 54 (34-81) | >99% (21597/21644) |
| Attending + APP | 1172 (20%) | Phys: 49 (30-76)<br>APP: 63 (39-110) | Phys: 48% (3846/7934)<br>APP: 51% (4077/7934) |
| Attending + Resident | 640 (11%) | Phys: 98 (63-138)<br>Resident: 129 (91-182) | Phys: 29% (1203/4217)<br>Resident: 71% (3007/4217) |

We also examined continuity of care across the included encounters. There was a median of 1(IQR 0-2) handoffs per encounter. Consistent with that, the number of identified frontline clinicians increased slowly with length of stay (Figure 3).

**Figure 3:**
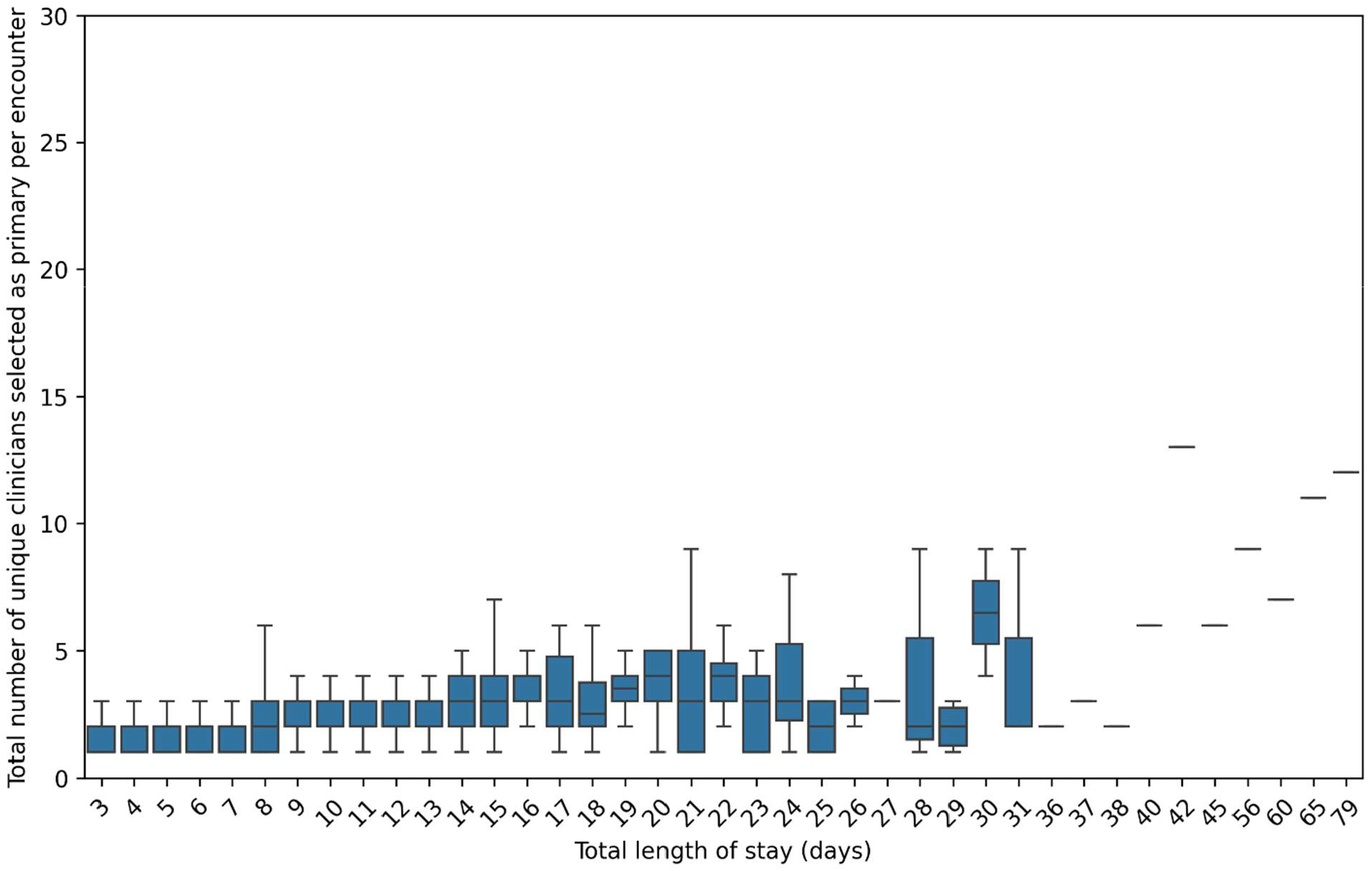
Number of distinct front-line physicians identified as a function of length of stay.

## Discussion

We developed an algorithm to identify the primary clinician for each patient-day using EHR audit log actions and clinician specialty data from login events and administrative data. Among the algorithmic variants, the hybrid algorithm that incorporated both specialty data sources (login and administrative data) achieved the highest accuracy relative to a chart review gold standard. In most misclassified cases, errors were driven by inaccuracies in data quality rather than by the audit log signal itself. Using the best performing approach, we found that most frontline clinicians in our hospital system were solo hospitalists, although care team models involving resident physicians and APPs were also present. There was reasonable continuity of care, with patients experiencing a median of 1 handoff during their inpatient stay.

Accurately identifying the clinician primarily responsible for a patient’s care is a long-standing measurement challenge, particularly in settings where care is delivered by teams and patients interacting with multiple clinicians within short time windows. Most prior attribution approaches have focused on the outpatient setting, and have relied on administrative claims or billing data, for example using majority or plurality (highest quantity) rules for number of physician visits or focusing on the largest amount billed by a clinician to identify primary care clinicians (3, 16–19). Validation studies have demonstrated the limitations of claims-based attribution, as agreement with gold-standard labels have ranged from 25-85% (16, 17).

Inpatient care introduces further attribution challenges, as claims-based approaches will struggle for care team models where the supervising clinician bills; the frontline clinician may not appear in the claims data. Visit- or cost-based selection rules also do not often translate to hospitalization-level accountability, motivating alternative inpatient attribution models that better reflect physician responsibility during a stay (12, 15, 18). Therefore, recent inpatient-focused work has begun integrating richer data sources to supplement claims-based methods for clinician attribution (2, 12, 15, 31–33), although the focus has largely been on incorporating note-writing or other documentation activities, which can be less reliable when clinicians share patient responsibility.

In this study, we leveraged EHR audit log data to infer responsibility at the patient-day level, as well as integrated specialty metadata to better disambiguate front-line clinician responsibility in team-based care. There are several advantages to our approach. First, audit logs provide a more holistic view of patient care for clinician attribution, as audit log actions encompass all EHR-related work, including documentation, chart review, and ordering activities. Notably, we found that while the clinician with the greatest number of audit log actions was usually the frontline clinician, consulting clinicians sometimes performed the most actions on the first day they evaluated the patient, so applying a specialty filter was important for separating the front-line clinician from consulting clinicians. Second, our approach is automated, scalable, and unobtrusive, as audit logs are automatically collected in the background without clinician intervention, thus avoiding the labor, cost, and potential Hawthorne effect associated with direct observation (20, 34). Third, our approach was designed for generalizability; it relied on total EHR actions rather than specialty- or role-specific activity markers, making it more applicable across roles and specialty types compared to methods that depend on metadata tailored to specific clinical contexts (2, 31–33). In addition, we focused specifically on inpatient clinicians, a group for whom attribution has historically been more challenging (15), and our algorithm showed the ability to attribute front-line clinicians even within supervisory care team structures (35).

Identification of frontline clinicians for inpatient medicine is important for outcomes research. Previous studies have found statistically significant associations between the physician responsible for inpatient care and patient outcomes, including mortality, quality of care, decisions to withdraw care, and hospice enrollment across a variety of clinical conditions (4, 5, 19, 36–38). In this context, primary clinician identity can function as an instrumental variable (capturing practice style, thresholds for escalation, or adherence to guidelines) or as an exposure of interest. Therefore, robust identification of front-line clinicians is an essential measurement component for studies focusing on quality improvement in clinical care delivery. Our approach could also be extended in future work towards attribution of other care team members, such as nurses, pharmacists, or other allied health professionals. Additionally, because audit logs are event-based and time-stamped, our approach naturally supports finer temporal resolution (e.g., shift- or hour-level attribution), which could be useful for describing teamwork during acute events. In addition to attribution, our approach also enables automated detection of handoffs in clinical care. We found that handoffs occurred a median of once per encounter, consistent with what has been found in other studies that have used scheduling or claims data to determine handoff frequency (28, 39), supporting the validity of our method. Scalable and accurate handoff detection is important because handoffs and discontinuity of care have been shown to be associated with longer lengths of stay, higher 30-day mortality, and higher hospital costs (7, 28, 40, 41).

### Limitations

This study had several limitations. First, although this study included 12 academic and community hospitals, all sites were affiliated with a single healthcare system. Therefore, generalizability to other healthcare systems and EHR platforms may be limited. Second, the algorithm focused on identifying a single primary clinician responsible for patient care on a given day. Although this worked well compared with manual chart review, it did struggle with collaborative care team structures where multiple clinicians shared responsibility for a patient. This care team structure was not common within our health system but may be more common elsewhere. Third, our method only captured EHR-mediated work and did not capture any work performed outside of the EHR. Therefore, we likely underestimated total work, although EHR work is likely correlated with total work. Finally, residual error was driven primarily by incorrect or inconsistent specialty labels rather than limitations of the audit log-based algorithm, highlighting that performance depends on the quality of institution-specific clinician metadata and may vary across sites.

## Conclusion

Our study demonstrated that audit log data, augmented with clinician specialty metadata, enabled reliable scalable identification of the primary front-line clinician for inpatient medicine encounters. The hybrid algorithm achieved high agreement with a chart review gold standard and produced patient-day assignments that could be used to quantify handoffs and continuity of care. Taken together, these findings provided a pragmatic, transportable framework for clinician attribution that could strengthen EHR-based teamwork, continuity of care, and outcomes research by improving how accountability and clinician-level effects are measured.

## Data Availability

All data produced in the present study are available upon reasonable request to the authors with appropriate data sharing agreements

## Clinical Relevance Statement

EHR audit log data, combined with clinician specialty data, can accurately identify the primary frontline inpatient clinician for each patient-day and has scalable potential. Our attribution method can strengthen inpatient outcomes and quality improvement studies by better linking care delivery and accountability to the clinicians providing day-to-day care.

## Protection of Human and Animal Subjects

This study was approved by the Washington University institutional review board with a waiver of informed consent (IRB no.: 202205084).

