## Supplemental tables 1-4 for "Development and validation of an algorithm to identify front-line clinicians using EHR audit log data"

**Supplemental Table 1:** Distribution of encounters selected for chart review across the inpatient hospitals affiliated with WashU Medicine and BJC Healthcare. Each location is further identified as being either academic or community locations. Values are reported as total counts and percentages of the chart review sample, n (%).

| **Hospital** | **Chart review subset (N=246)** | **Entire cohort (N=5801)** |
|---|---|---|
| Location 1 (community) | 23 (9%) | 546 (9%) |
| Location 2 (academic) | 48 (20%) | 1118 (19%) |
| Location 3 (community) | 15 (6%) | 363 (6%) |
| Location 4 (community) | 6 (2%) | 149 (3%) |
| Location 5 (community) | 37 (15%) | 863 (15%) |
| Location 6 (community) | 31 (13%) | 725 (12%) |
| Location 7 (community) | 5 (2%) | 118 (2%) |
| Location 8 (community) | 40 (16%) | 949 (16%) |
| Location 9 (community) | 21 (9%) | 503 (9%) |
| Location 10 (community) | 1 (0.4%) | 2 (0.03%) |
| Location 11 (community) | 12 (5%) | 297 (5%) |
| Location 12 (community) | 7 (3%) | 168 (3%) |

**Supplemental Table 2:** Pairwise comparisons among the four algorithms were performed using McNemar's tests; p-values were Bonferroni corrected to account for multiple comparisons and capped at a value of 1.0.

| Pairwise comparison | P value raw | P value corrected |
|---|---|---|
| Algorithms 1 vs 2 | 5.7e-18 | 3.4e-17 |
| Algorithms 1 vs 3 | 0.0015 | 0.0091 |
| Algorithms 1 vs 4 | 1.0e-27 | 6.3e-27 |
| Algorithms 2 vs 3 | 3.7e-9 | 2.2e-8 |
| Algorithms 2 vs 4 | 1.0e-3 | 6.2e-3 |
| Algorithms 3 vs 4 | 3.2e-20 | 1.9e-19 |

**Supplemental Table 3:** The distributions of supervision models stratified by hospital among all encounters. Supervision models are categorized as physician alone – no supervision structure, physician + APP - physician supervising an APP, or physician + resident- physician supervising a resident. Data is presented as total counts (%), median (IQR) actions per day, and fraction of days (%) each role performed audit log actions.

| Supervision structure | Number of Encounters | EHR actions per day, median (IQR) | Fraction of days with each role present |
|---|---|---|---|
| **Location 1** | | | |
| Physician alone | 344 | 62 (44-87) | Phys: 99.9% (1870/1877) |
| Physician + APP | 103 | Phys: 47 (29-76)<br>APP: 80 (52-112) | Phys: 40% (254/638)<br>APP: 60% (382/638) |
| Physician + Resident | 99 | Phys: 89 (60-116)<br>Resident: 115 (78-158) | Phys: 33% (171/524)<br>Resident: 67% (351/524) |
| **Location 2** | | | |
| Physician alone | 391 | 82 (48-126) | Phys: 99.9% (2327/2328) |
| Physician + APP | 162 | Phys: 83 (48-130)<br>APP: 152 (104-216) | Phys: 43% (491/1142)<br>APP: 56% (650/1142) |
| Physician + Resident | 541 | Phys: 100 (64-141)<br>Resident: 131 (93-185) | Phys: 28% (1032/3693)<br>Resident: 72% (2656/3693) |
| **Location 3** | | | |
| Physician alone | 185 | 51 (35-72) | Phys: 99% (932/934) |
| Physician + APP | 178 | Phys: 44 (29-64)<br>APP: 52 (39-73) | Phys: 27% (276/1010)<br>APP: 72% (732/1010) |
| **Location 4** | | | |
| Physician alone | 96 | 120 (83-164) | Phys: 99% (444/445) |

| | | | |
|---|---|---|---|
| Physician + APP | 53 | Phys: 111 (72-170)<br>APP: 130 (85-233) | Phys: 57% (157/275)<br>APP: 43% (118/275) |
| **Location 5** | | | |
| Physician alone | 607 | 46 (30-71) | Phys: 99% (3122/3132) |
| Physician + APP | 256 | Phys: 39 (25-60)<br>APP: 43 (27-64) | Phys: 45% (984/2206)<br>APP: 55% (1218/2206) |
| **Location 6** | | | |
| Physician alone | 686 | 51 (34-75) | Phys: 99% (3827/3830) |
| Physician + APP | 39 | Phys: 45 (31-67)<br>APP: 81 (52-113) | Phys: 82% (222/270)<br>APP: 17% (46/270) |
| **Location 7** | | | |
| Physician alone | 68 | 42 (24-59) | Phys: 100% (294/294) |
| Physician + APP | 50 | Phys: 33 (20-56)<br>APP: 9 (2-20) | Phys: 70% (202/287)<br>APP: 30% (85/287) |
| **Location 8** | | | |
| Physician alone | 859 | 47 (30-71) | Phys: 99% (5054/5067) |
| Physician + APP | 90 | Phys: 50 (34-69)<br>APP: 56 (35-87) | Phys: 85% (602/709)<br>APP: 15% (107/709) |
| **Location 9** | | | |
| Physician alone | 404 | 53 (37-75) | Phys: 99% (2186/2192) |
| Physician + APP | 99 | Phys: 51 (38-73)<br>APP: 78 (54-266) | Phys: 70% (406/580)<br>APP: 30% (174/580) |
| **Location 10** | | | |

| | | | |
|---|---|---|---|
| Physician alone | 2 | 42 (12-55) | Phys: 100% (9/9) |
| **Location 11** | | | |
| Physician alone | 244 | 65 (44-91) | Phys: 99% (1188/1192) |
| Physician + APP | 53 | Phys: 56 (37-84)<br>APP: 103 (73-127) | Phys: 29% (90/306)<br>APP: 70% (216/306) |
| **Location 12** | | | |
| Physician alone | 79 | 38 (25-58) | Phys: 100% (344/344) |
| Physician + APP | 89 | Phys: 42 (25-59)<br>APP: 53 (33-76) | Phys: 32% (162/511)<br>APP: 68% (349/511) |

**Supplemental Table 4:** The distributions of supervision models stratified by hospital among the 246 chart reviewed encounters. Supervision models are categorized as physician alone – no supervision structure, physician + APP - physician supervising an APP, or physician + resident-physician supervising a resident. Data is presented as total counts (%), median (IQR) actions per day, and fraction of days (%) each role performed audit log actions.

| Supervision structure | Number of Encounters | EHR actions per day, median (IQR) | Fraction of days with each role present |
|---|---|---|---|
| **Location 1** | | | |
| Physician alone | 18 | 64 (42-84) | Phys: 100% (89/89) |
| Physician + APP | 2 | Phys: 92 (92-92)<br>APP: 86 (53-114) | Phys: 17% (1/6)<br>APP: 83% (5/6) |
| Physician + Resident | 3 | Phys: 90 (90-90)<br>Resident: 74 (56-92) | Phys: 7% (1/14)<br>Resident: 93% (13/14) |
| **Location 2** | | | |
| Physician alone | 19 | 91 (61-142) | Phys: 100% (93/93) |
| Physician + APP | 4 | Phys: 128 (100-177)<br>APP: 145 (74-209) | Phys: 18% (3/17)<br>APP: 82% (14/17) |
| Physician + Resident | 21 | Phys: 75 (64-120)<br>Resident: 127 (96-169) | Phys: 24% (34/143)<br>Resident: 76% (109/143) |
| **Location 3** | | | |
| Physician alone | 5 | 46 (34-53) | Phys: 100% (21/21) |
| Physician + APP | 10 | Phys: 63 (42-91)<br>APP: 53 (34-73) | Phys: 6% (3/53)<br>APP: 69% (50/53) |
| **Location 4** | | | |
| Physician alone | 5 | 114 (68-143) | Phys: 100% (22/22) |
| Physician + APP | 1 | Phys: 88 (69-177)<br>APP: 61 (61-61) | Phys: 75% (3/4)<br>APP: 25% (1/4) |
| **Location 5** | | | |

| | | | |
|---|---|---|---|
| Physician alone | 31 | 43 (26-68) | Phys: 100% (160/160) |
| Physician + APP | 6 | Phys: 41 (29-50)<br>APP: 57 (34-76) | Phys: 17% (5/30)<br>APP: 83% (25/30) |
| **Location 6** | | | |
| Physician alone | 31 | 48 (31-73) | Phys: 100% (155/155) |
| **Location 7** | | | |
| Physician alone | 5 | 38 (25-42) | Phys: 100% (18/18) |
| **Location 8** | | | |
| Physician alone | 40 | 51 (29-75) | Phys: 100% (242/242) |
| **Location 9** | | | |
| Physician alone | 18 | 67 (44-84) | Phys: 100% (102/102) |
| Physician + APP | 3 | Phys: 55 (44-68)<br>APP: 53 (51-80) | Phys: 70% (14/20)<br>APP: 30% (6/20) |
| **Location 10** | | | |
| Physician alone | 1 | 76 (64-88) | Phys: 100% (2/2) |
| **Location 11** | | | |
| Physician alone | 10 | 70 (53-104) | Phys: 100% (44/44) |
| Physician + APP | 2 | Phys: 79 (79-79)<br>APP: 138 (88-160) | Phys: 13% (1/8)<br>APP: 88% (7/8) |
| **Location 12** | | | |
| Physician alone | 3 | 49 (39-73) | Phys: 100% (10/10) |
| Physician + APP | 4 | Phys: 39 (25-56)<br>APP: 76 (61-89) | Phys: 27% (4/15)<br>APP: 73% (11/15) |
